# Stimulating the Motor Development of Very Premature Infants: Effects of Early Crawling Training on a Mini-Skateboard

**DOI:** 10.1101/2023.03.24.23287528

**Authors:** Marie-Victorine Dumuids-Vernet, Vincent Forma, Joëlle Provasi, David Ian Anderson, Elodie Hinnekens, Evelyne Soyez, Mathilde Strassel, Léa Guéret, Charlotte Hym, Viviane Huet, Lionel Granjon, Lucie Calamy, Gilles Dassieu, Laurence Boujenah, Camille Dollat, Valérie Biran, Marianne Barbu-Roth

## Abstract

**Aim:** To examine the effects of an early home-based 8-week crawling intervention performed by trained therapists on the motor and general development of very premature infants during the first year of life.

**Methods:** At term-equivalent age, immediately following discharge from the Neonatal Intensive Care Unit (NICU), we randomly allocated 44 premature infants born before 32 weeks’ gestation without major brain damage to one of three conditions in our intervention study: crawling on a mini-skateboard, the Crawliskate (Crawli), prone positioning control (Mattress), or standard care (Control). The Crawli and Mattress groups received 5 min daily at-home training administered by trained therapists for 8 consecutive weeks upon discharge from the NICU. The outcomes of greatest interest included gross motor development (Bayley-III) at 2, 6, 9, and 12 months (primary outcome) corrected age (CA), mature crawling at 9 months CA and general development at 9 and 12 months CA [Ages and Stages Questionnaires-3 (ASQ-3)].

**Results:** A 3 (Condition) x 4 (Age) repeated measures ANOVA revealed that Crawli group infants had significantly higher Bayley-III gross motor development scores than Mattress and Control group infants. Crawli group infants also scored significantly higher on groups of Bayley-III items related to specific motor skills than infants in the other groups, including crawling at 9 months CA (p<0.05). Separate one-way ANOVAs at each of the four ages tested revealed the Crawli group had significantly higher gross motor development scores than the Mattress group (p<.001) but not the Control group (p<.08) at 2 months CA, than the Mattress and Control groups at 6 months CA (p<.05), and than the Control group at 9 and 12 months CA (p<.05). The Mattress and Control groups did not differ significantly at any age. A 3 (Condition) x 2 (Age) repeated measures ANOVA revealed that the Crawli group scored significantly higher than the Control group for the ASQ-3 total score (p<.01) and communication score (p<.05) and significantly higher for the fine motor score than the Control (p<.01) and Mattress (p<.05) groups. We found additional significant differences in favor of the Crawli group for other dimensions of the ASQ-3 in separate one-way ANOVAs at 9 and 12 months CA. **Interpretation** Early crawling training on a Crawliskate provides an effective way to promote motor and general development in very premature infants. The Crawli group’s significantly higher scores on the crawling items at 9 months CA provide clear evidence for a link between newborn crawling and more mature crawling later in development.

**What this paper adds:**

- Very premature infants can propel themselves on a mini-skateboard using crawling movements at term equivalent age
- Eight weeks of daily, at-home early crawling training immediately following discharge from the NICU facilitates the acquisition of mature crawling in premature infants
- Eight weeks of early crawling training positively influences motor development in premature infants
- Eight weeks of early crawling training positively influences general development in premature infants
- Early daily at-home crawling is a promising intervention for premature infants at heightened risk for motor delays and disabilities, potentially feasible for parents to conduct

**Data sharing statement:** The data that support the findings of this study are available on request from the corresponding author. The data are not publicly available due to privacy or ethical restrictions.

## INTRODUCTION

Preterm birth, defined as delivery prior to 37 weeks gestational age (WGA), is a worldwide epidemic with a world global incidence of approximately 15 million preterm births per year (1). Prematurity continues to increase at a constant rate even though it appears to have stabilized recently in some countries (2). In parallel, the survival rate of children born prematurely has increased, particularly for very premature babies born before 32 WGA (around 20% of premature births), thanks to advances in medical practices and the quality of neonatal services. Paradoxically, this situation creates a real public health problem because the increase in the survival of very premature infants leads to an increase in children with disabilities during their development. Yearly, 5-8% of very preterm born survivors develop cerebral palsy (CP), which represents the main cause of childhood disability (3,4), resulting in significant delays and/or impairments in postural, manual and locomotor development, depending on the severity of the CP. Even if they do not develop CP, very premature babies remain at risk for motor problems, with 40% presenting developmental delays or impairments, including later sensorimotor anomalies that can impact gross motor development (5).

### The centrality of motor/locomotor development in human development

Given the fundamental role movement plays in all human behavior, delays and impairments in the acquisition of motor skills during the first year of life have significant implications for the subsequent development of the brain and behavior and for an individual’s ultimate quality of life. Researchers have established that the acquisition of motor skills during the first year of life has cascading effects on the later development of skills in the motor and psychological domains in at-term and preterm infants (6–13). Several studies have shown that the quality of early movements in preterm infants predict the quality of motor skills at later ages (12,14,15). Hua and colleagues recently showed that even a mild delay in crawling and walking onsets increases the risk for subsequent motor impairments in childhood (16). Research on the psychological revolutions that follow the acquisition of independent crawling and walking provides some of the strongest evidence for the fundamental contribution motor development makes to broader developmental outcomes (6,7,17–19). Researchers have linked the emergence and practice of independent crawling around 8-9 months of age to changes in perceptual-action coupling, spatial cognition, memory, social and emotional functioning and brain functioning (6,17,20– 25) and the acquisition of walking to a range of developmental changes (26), particularly in the language domain (8,27,28).

The pervasive effect of locomotor experience on a child’s development represents one of the primary reasons clinicians target locomotor skills for therapeutic intervention for children at risk for developmental delay. However, most of the interventions designed to promote locomotion begin well after the age of 12 months, largely because clinicians cannot diagnose locomotor problems and neurological disorders like CP until infants have already demonstrated delayed acquisition of motor skills. A recent systematic review by Dumuids-Vernet and collaborators reported that since the year 2000, only ten motor/locomotor interventions commenced before the age of one year in infants at risk for motor delays and only three targeted infants born preterm, the other studies targeted infants with an established disability at birth (Myelomeningocele, Down syndrome or high brain damage) (29).

### The need for early motor/locomotor interventions for very premature infants

Given the heightened risk of motor/locomotor delays and impairments in very preterm infants, it is essential to design early intervention programs that clinicians could start even before the diagnosis of a behavioral or neurological disorder to improve long-term motor outcomes for these infants (30,31). As indicated by a growing body of literature (30,32) these programs should be implemented as early as possible to take advantage of the heightened plasticity of body and supra spinal structures (i.e., brain and corticospinal tract) and spinal circuitries (33–35). It is also essential (30) that these programs: 1) promote active, self-generated movements, 2) are high in frequency (30,31,35–37), and 3) target specific functions that are likely to generalize more broadly to motor and muscular development and be feasible for infants limited in their postural control and mobility (29,38). What type of very early intervention could be implemented to stimulate active locomotion in very premature infants while adhering to these recommendations? Could an intervention designed to stimulate locomotor development be started as early as term equivalent age and would such an intervention facilitate the development of mature locomotion and other gross motor skills?

### The link between primitive neonatal locomotion and mature crawling and walking

Human newborns can perform alternating locomotor movements of their legs at birth when supported upright in a stepping position on a table (39,40) and can propel themselves forward when breast crawling on their mother’s abdomen (41,42) and via swimming movements when supported in the prone position in water (43). Researchers and clinicians have observed these patterns in preterm infants in neonatal care units when they crawl occasionally in their incubators (44) and when they perform stepping movements while supported upright under their armpits (45,46). Though traditionally considered simple spinal reflexes, destined to rapidly disappear under the increasing influence of cortical maturation and playing no role in the development of independent locomotion, a growing body of literature has revealed that these primitive locomotor behaviors display much greater complexity than simple reflexes and serve as important precursors to mature crawling and walking. First, primitive locomotion does not disappear during development; biomechanical constraints mask its expression. For example, a rapid increase in fat mass during the first two months of life make the legs difficult to lift in the upright position if the infant has not received stepping practice, leading to an apparent disappearance of the stepping pattern. However, minimizing the gravitational force on the legs by submerging the infant in water (47,48) or driving leg extension with a moving treadmill belt (49) can reactivate the stepping pattern between 2 and 6 months of age.

The second reason researchers now consider early motor patterns as complex behaviors is that several studies have shown that a range of higher order stimuli processed supra spinally can initiate and modulate newborn stepping and crawling, including visual, olfactory, and auditory stimuli (50–54). Finally, researchers have used a variety of experimental approaches to document a link between primitive and mature walking. One approach has used EMG recordings to show common patterns of muscle activation in newborn stepping and mature walking (55), even though such patterns remain plastic during development and can be fractioned into more controlling units (56). In another approach, researchers have shown that two months of daily stepping on a solid surface from birth leads to an earlier emergence and/or higher quality of mature walking in typically-developing infants (57,58) and training stepping on a treadmill leads to an earlier emergence and higher quality of walking in infants with Down syndrome (59–63). Though less studied, researchers have shown that daily training of supported crawling in typically developing infants from four months of age onward leads not only to an earlier emergence of independent crawling and walking but boosts cognitive development (19). In summary, a large body of literature provides evidence for three important linkages in early development, each with connections to the other linkages. First, infants born very preterm have a much greater risk for delayed and/or impaired motor development, including locomotor development. Second, early primitive locomotor patterns appear to serve as precursors for later emerging independent crawling and walking. Third, the acquisition of crawling and walking make fundamental contributions to the development of a diversity of skills in the motor and psychological domains. Taken together, these linkages suggest that training locomotion in infants born very preterm could have positive and pervasive effects on their developmental outcomes. This suggestion raises some interesting questions. For example, what type of active locomotor movements should clinicians stimulate? Should the infant’s movements be active or passive? When should the stimulation begin? Is there a preferred method or paradigm for providing the stimulation? Researchers have used a solid surface or treadmill to promote stepping in young infants, however stepping on these surfaces could create problems for very preterm infants, who are more fragile and often hypo or hypertonic. Furthermore, this type of stepping practice requires an experimenter to support the infant’s body and head and constrains independent movement and forward propulsion. Moreover, supporting the infant’s head and body under the armpits blocks potential locomotor movements of the arms and associated trunk and head movements, important in mature crawling and walking (64,65).

### The potential benefits of stimulating quadrupedal locomotion early in life

The stimulation of quadrupedal locomotion has several advantages over the stimulation of other motor activities, such as isolated head movements, stepping on a treadmill, or repetitive limb movements. Crawling represents a whole-body activity that involves the coordination and sequencing of multiple muscles and joints, leading to observable propulsion and reorientation of the infant to her environment, even in the newborn. Notably, the neuromuscular substrate underlying the quadrupedal organization of crawling also seems to underlie bipedal stepping and walking (64,65). Consequently, the active practice of crawling potentially facilitates the development of not only quadrupedal locomotion but also bipedal locomotion, holding of the head, unsupported sitting (by strengthening the muscles of the neck and trunk) and standing. However, due to the weight of their head, even typically developing newborns cannot lift their trunk and head to move forward by themselves in a prone position at birth; an experimenter must support very young infants (45,46,66), preventing them from moving actively in any direction in their environment.

### The Crawliskate

Given the aforementioned limitations in newborns’ ability to propel themselves via crawling movements, we designed and constructed a mini-skateboard, which raises the head and trunk off the ground and allows the arms to move freely, to facilitate and stimulate quadrupedal locomotion during the first months of life (67). This mini-skateboard, known as the Crawliskate, promotes a lengthening of the spine and a neutral position of the head to facilitate the arm and leg movements necessary to generate independent propulsion (see a detailed description in the methods). The results of a previous study on 60 typically-developing newborns highlighted the advantages of using the Crawliskate to promote newborn crawling (67). The newborns propelled themselves significantly further on the Crawliskate and they demonstrated more mature crawling patterns, in terms of limb kinematics and interlimb coordination, than when observed crawling without the Crawliskate.

This current study investigates the feasibility and the effect of early crawling training using the Crawliskate on the locomotor and motor development of very preterm infants. The intervention complies with the essential program elements for early interventions described earlier in the introduction, particularly for infants with limited postural control or who are hypotonic, and it preserves the natural alignment of the head, neck and spine when the infant is in the prone position, which is particularly important for infants born premature. The “Crawliskate” can be used at even younger ages than a robotic device recently designed to train crawling in infants with or at risk for cerebral palsy (68,69) and it has the added benefits of being portable and easy to use in the home. As noted earlier, studies of more than four hundred typically-developing newborns revealed that neonatal crawling can be regulated by visual, olfactory and auditory stimuli processed supra spinally, and it appears to engage and stimulate the neural circuitry underlying the control of mature bipedal and quadrupedal locomotion (53,54,67). Consequently, newborn crawling represents an ideal target for early intervention. Early crawling training has the potential to influence the plasticity and development of the supra spinal tracts, ultimately contributing to the development of not only locomotion but a range of gross motor skills and the cognitive and psychological skills to which these motor skills have been linked (6,17,19).

### Objectives

Given the aforementioned observations of typically-developing newborns, we hypothesized that stimulating very premature infants’ crawling on the Crawliskate would enhance their mature crawling (and possibly their walking), gross motor, and general development during the first year of life. We based our expectations on the findings from studies referred to earlier showing that five minutes of daily upright stepping practice over the first eight weeks of life accelerates walking onset of typically developing infants (57,58), presumably because of the well-established neurological links between newborn stepping and later walking (55), and 15 minutes of daily crawling training in 5-9 month-old typically developing infants accelerates crawling onset, motor development and intellectual development relative to controls (19). We anticipated these latter training effects would be more pronounced in very young infants given their heightened body and brain plasticity (34,35). The long-term aim of the study was to compare the motor and general development of infants up to 60 months corrected age (CA), however, in the current paper we present the motor and general outcomes up to 12 months CA.

## METHOD

### 1. Trial design

The study was registered at www.clinicaltrials.gov; registration number: NCT05278286. Because of the limited number of subjects in our study due to the home-based nature of our intervention (see training protocol below), we chose not to include very preterm infants with major brain lesions because this would have required us to match lesion levels and locations across the three groups and we could not test a large enough sample to accomplish such matching. We randomly allocated forty-four very preterm infants without major brain lesions to one of three groups: Crawling training on the Crawliskate (Crawli), prone positioning on a Mattress training without the Crawliskate (Mattress), or standard care (Control), in which infants received pediatric checkups at 4, 9, 12, 18 and 24 months CA but no other treatment unless recommended by the pediatrician. It is important to note that we designed the Mattress condition to act as an active Hawthorne-effect control for the effect of prone positioning on the Crawliskate (prone positioning has positive effects on motor development, e.g. (70)) and the effect of visits from the osteopaths who implemented the training in the infants’ homes. Because infants born between 24+0 and 27+6 (GA+Days) have higher risk for developmental delays than those born between 28+0 and 32+0 GA (71), the randomization process stratified infants on gestational age (24-27+6 GA vs 28-32 GA) to ensure equal rates of GA at term equivalent age between groups.

### 2. Participants

We recruited eligible infants between March 2017 and October 2019 from four Neonatal Intensive Care Units (NICU) in Paris, France. Infants were included if they were born between 24 and 32 weeks of gestation (WG), lived within 10 kms of our laboratory (because of daily home visits by our team for the training groups), and could leave the NICU and begin training between 37 and 42 WG. Exclusion criteria included congenital anomalies, major brain damage (such as IVH3 and 4) defined by transcranial ultrasound, bronchopulmonary dysplasia with oxygen dependence after 36 WG, any digestive or other problems preventing prone positioning, limb deformities, and presence of retinopathy or sensory pathology that may delay motor development. We excluded infants if only one of the criteria was present. The study was approved by the ethical committee Ile de France 3 (CPP n°3465; ANSM n°2016-A01320-51). All parents provided written informed consent for their infants to participate in the study.

### 3. Detailed description of the Crawliskate

The Crawliskate, or Crawli, is a mini skateboard, adapted to newborns and young infants, that permits comfortable positioning on the belly. The slight upward slope (15 degrees) of the Crawli allows the upper part of the trunk and the head to be elevated, with the head resting on a platform in front (Figure 1D). The slope also allows maintenance of the natural flexion of the trunk and head, avoiding the possible hyper-extension of the head that can occur when the infant is placed directly in a prone position on a flat surface. The other advantage of the Crawli is that it frees the movements of the arms because the upper trunk is raised. Under these conditions, the arms can actively participate in the child’s propulsion along with the legs (67). With the minimum slope of the Crawli at the infant’s pelvis, the infant’s legs rest on the surface of support, which allows the infant to learn to elevate the pelvis to make effective propulsive thrusts. To ensure the safety of the infant, he/she is secured to the Crawli by a system of fabric fins and straps that wrap the child on the skate (Figure 1A & 1B). Two stabilizers placed on each side and in front of the headrest prevent any lateral imbalance during the infant’s propulsive pushes. The wheels placed under the Crawli allow the infant to move 360 degrees.

**Figure 1.**
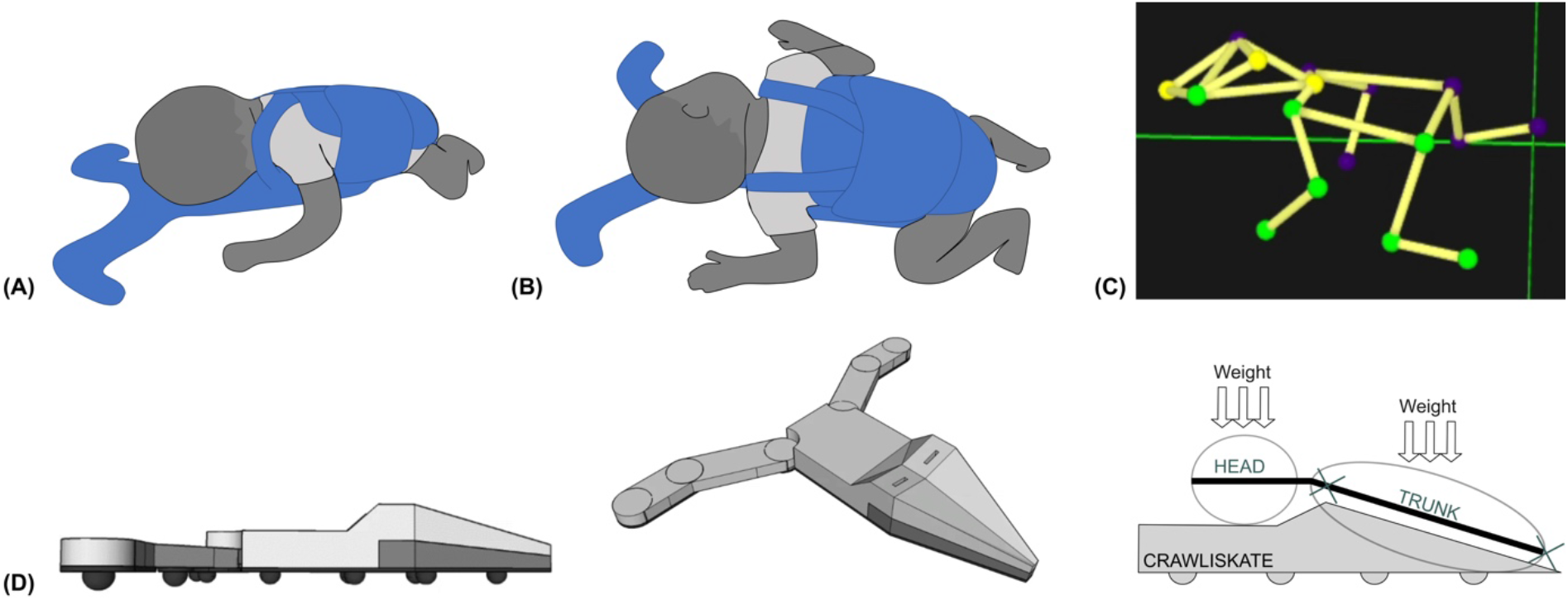
(A) A newborn resting on its belly and wrapped on the Crawli; **(B)** Quadrupedal propulsion on the Crawli; **(C)** 3D reconstruction of quadrupedal crawling on the Crawli; **(D)** Structure of the Crawli. Patent: Barbu-Roth, M., Forma, V., Teulier, C., Anderson, D., Provasi, J., & Schaal, B. (2016). -Device for assisting the crawling of an infant. Patent WO2016009022. https://patentscope.wipo.int/search/en/detail.jsf?docId=WO2016009022

The Crawli thus allows the infant to move actively in any direction by pushing with all four limbs (Figure 1B & 1C for a newborn on the Crawliskate).

### 4. Intervention at home

#### 4.1. Training of the osteopaths in charge of the Crawli and Mattress training at home

We recruited thirty professionals to conduct the intervention. Before they implemented the intervention, Evelyne Soyez, a senior physiotherapist, expert in neurodevelopmental disorders, trained them. This training included learning Albert Grenier’s method for handling fragile infants (72), an introduction to the issue of neurodevelopmental disorders, and an introduction to Claudine Amiel-Tison’s neurological examination: ATNAT (73). Each osteopath also had to complete a Red Cross training course and earn a diploma in pediatric emergency care (IPSEN training, Initiation Premier Secours Enfants Nourrissons).

Then, the lead osteopath on our team, Marie-Victorine Dumuids-Vernet, who organized all training sessions for this study, taught the 30 osteopaths the protocol for the home visits and the training for the infants in the Crawli and Mattress training groups. The lead osteopath conducted the training initially in a classroom at our laboratory using a baby doll and the Crawliskate and then at home with the infant and parents. The lead osteopath also supervised each of the other osteopaths as they conducted the first seven days of training in the home. We monitored the quality of the training implemented by each osteopath by viewing daily videos of the training sessions at the end of each week of training. This standardized protocol allowed us to ensure the homogeneity of the trainings across the different trainers.

#### 4.2. Training protocol

Each infant included in the Crawli and Mattress groups received 5 minutes of daily training for 8 consecutive weeks at home by an osteopath. The training started as soon as the infant was between 37 and 42 WG and left the NICU. The five minutes duration of crawling training on the Crawli is consistent with a study that trained upright stepping (58) and similar to a study that used a robotic device to train crawling in older infants (68,69).

##### 4.2.1. Crawli training

The osteopath gently placed the infant on the Crawli, with the head at the level of the clean bib on the headrest platform, and so the infant’s hands and feet contacted a thin 1meter x 2 meters mat beneath the device. The infants propelled themselves independently on the device. During the session, particularly during the first week of training, the osteopath occasionally placed a hand behind the infant’s foot or hand to provide a surface to push against and/or extended the fingers of the infant’s hand to facilitate pushing and teach the infant how to place her feet and hands to push effectively and efficiently (see Figure 2A^1^ and the beginning of the video Crawli Home). The osteopath was careful not to create any forward propulsion, as the infant should be the only one actively initiating propulsion (see the videos Crawli Home or Crawli Lab for a full independent crawling of the infant on the Crawli). During the five minutes of training, the caregiver could initiate a break if the infant became too agitated or regurgitated.

**Figure 2.**
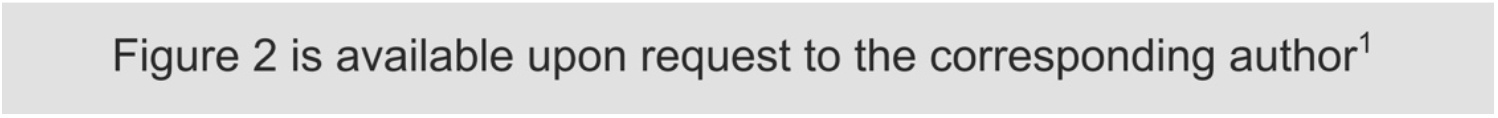
(A) Photo of the beginning of a Crawli training session at home with the osteopath assisting placement of the feet of the infant on the mat. **(B)** Photo of a Mattress training session at home.

##### 4.2.2. Mattress training

The mattress training was almost identical to the Crawliskate training, except that the infant was placed directly on his belly on the mat (Figure 2B^1^ and video Mat Home^1^) and the osteopaths did not use their hands to provide a surface for the infants to push against because pushing this way while the friction from the surface blocked the infant’s forward progression could have increased the stress on the infant’s neck to a harmful level. With the infant lying prone on the mat, the osteopath allowed the infant to make his or her own movements, taking care to monitor the upper airway clearance. Again, breaks were provided if the infant became too agitated or regurgitated and the session could be shortened if the infant could not be calmed down or showed signs of fatigue.

It is important to note that the parents played no role in training their infants other than to be present to calm their infants if they experienced distress. The osteopaths conducted all of the training. To ensure that parents did not practice training outside of the sessions performed by our team, we stored the training material (Crawliskate, mats, cameras) in a locked bag stored at the parents’ home.

#### 4.3. Safety monitoring

##### 4.3.1. Before training

To ensure infants tolerated prone positioning, we performed a one-minute test with each infant on the Crawliskate or without the Crawliskate but prone on the mat before training started. During this test, we monitored the infant’s oxygen saturation and heart rate while observing their behavior and checking how well they tolerated the position (see videos Crawli Lab^1^ and Mat Lab^1^ to see infants tested at term equivalent age at our babylab). All infants tolerated the prone position on the Crawliskate or the mat.

##### 4.3.2. During training

Diaries kept by the osteopaths and videos of the home trainings allowed us to check for any adverse events. We observed no adverse events.

##### 4.3.3. After training

Each infant was followed by our senior physical therapist and osteopath, Evelyne Soyez, who assessed the infants using the ATNAT up to 12 months CA and discussed with the parents any concerns they had about their infants’ development after 2 months CA (the senior physical therapist was blind to the infant’s group). Parents were also asked to report activities they engaged in with their infants when their infants were 2, 6, 9, and 12 months CA, which was designed to track how the infants were stimulated after the training sessions. Neither the osteopaths, parents, nor the experimenters observed or reported any adverse events.

#### 4.4. Recording of training and evaluation of traveled distance

All training sessions at home were videotaped with two GoPros cameras placed on each side of the mat to ensure the osteopaths implemented the training correctly and to measure distances traveled by the infants at each session. We measured the traveled distance covered by the infant at each session in cm by coding the videos of each session. We compared the mean traveled distances of the infants in the Crawli and Mattress groups to evaluate how well they could propel themselves as a function of the training protocol. We also charted the progression over time in the average distances covered each week between the first and last training session for the Crawli group (the Mattress group showed very limited propulsion – see the results section).

### 5. Outcomes

Figure 3 summarizes the outcome assessments. The assessors of all outcomes were blind to the infants’ group and the osteopaths who performed the training had no contact with the assessors. Note, we do not present the crawling efficiency data (assessed via three-dimensional kinematics) at term equivalent age, 2, 6, 9, and 12 months CA in this manuscript because we continue to reduce and analyze those data.

**Figure 3.**
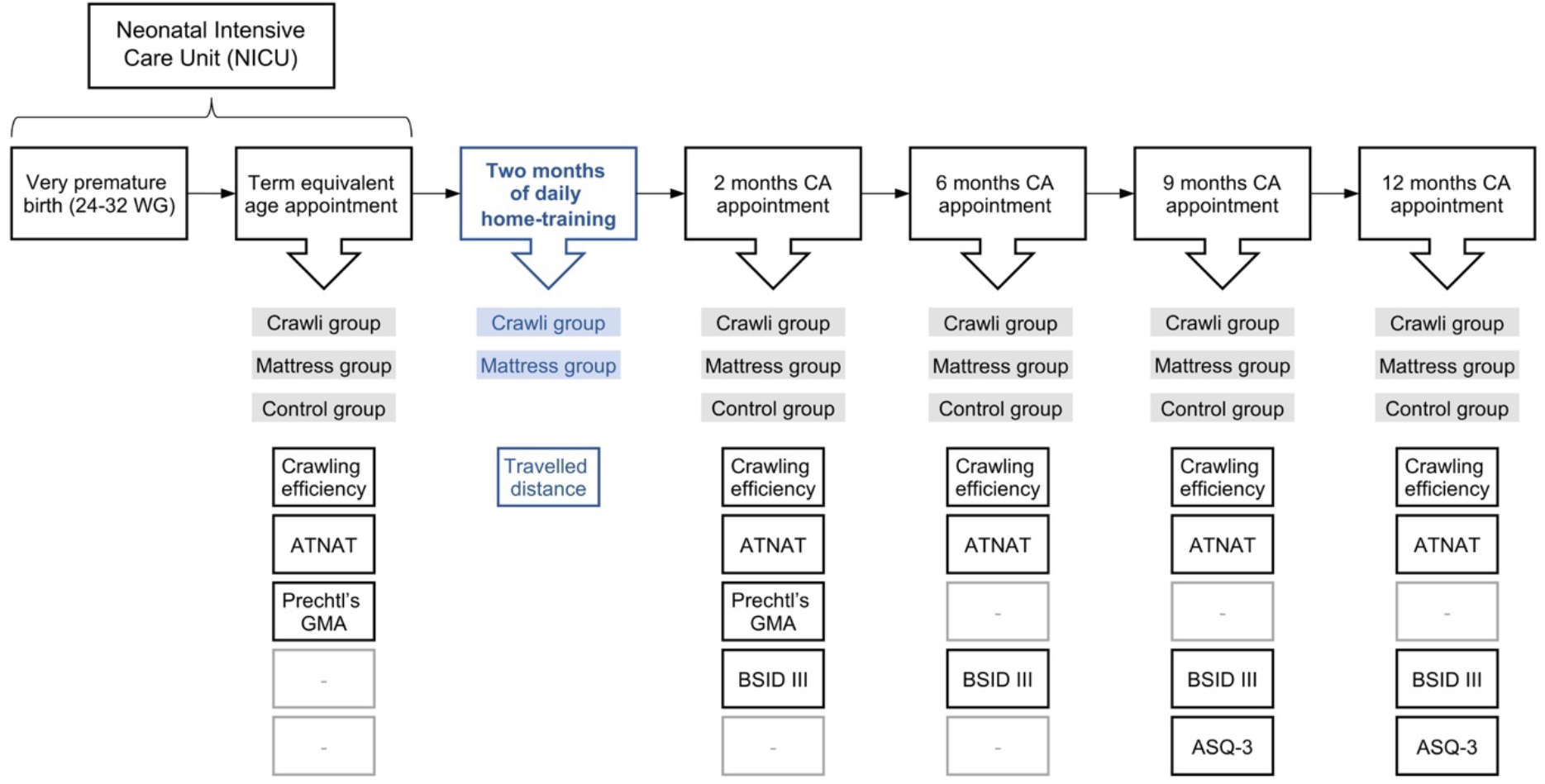
Premalocom first year follow-up.

#### 5.1. Gross and Fine Motor Bayley Assessments

We administered the Bayley III gross and fine motor assessments at 2, 6, 9, 12, and a full Bayley III assessment at 24 months CA (BSID-III ed. (74) Bayley, 2006) to assess general motor development and to evaluate the development of specific motor milestones, especially mature crawling, but also head control, sitting, standing and walking. Two trained testers administered the assessments. They both had over 20 years of experience assessing infant development and 10 and 8 years of experience administering the Bayley assessments. Both were blinded to infants’ group assignment. Scaled scores were calculated. Before the start of the study the raters assessed 8 infants between 2 and 12 months of age using the BSID-III and were required to reach a high interrater reliability [intra class correlation coefficient (ICC) > 0.9]. During the study, 10% of the assessments were double scored by the two raters in real time and all assessments were videotaped so disagreements could be resolved afterward. The interrater reliability was excellent (ICC = 0.92).

#### 5.2. ASQ-3 General Assessment

We asked parents to complete the ASQ-3 questionnaire to evaluate their infants’ general development in the communication, gross motor, fine motor, problem solving, and personal-social domains at 9, 12, 18, 24 and 60 months CA^2^, as performed in previous studies on premature infants (4,5). We chose to administer the ASQ-3 from 9 months of age onward for two reasons. First, we did not want to overwhelm the parents at the beginning of the study by making too many demands on them. Second, we expected differences between groups to appear in the general domains assessed by the ASQ-3 after 8-9 months of age, when typically-developing infants generally acquire the ability to crawl independently. We based our expectations on the range of psychological changes researchers have documented following the onset of independent crawling.

#### 5.3. Amiel-Tison Neurological Assessment (ATNAT)

The ATNAT clinical examination assesses the quality of musculoskeletal and neuromotor function responses to tonic and active stimulation (75). ATNAT scores were evaluated at term equivalent age and at 2, 6, 9 and 12 months CA by our senior physiotherapist, Evelyne Soyez, with 30 years of experience administering the ATNAT to ensure the groups were equivalent at the beginning of the study, to monitor for any deleterious effects of the training, and to diagnose any infants at heightened risk for a neurological disorder. The specialist was blinded to the infants’ group assignment.

#### 5.4. Prechtl’s General Movement Assessment (GMA)

The GMA examines the quality of the spontaneous movements of the infant while she is lying supine on a flat surface in a calm but alert state, stage 3 (76), without any external stimulation. The GMA was performed at term equivalent age and at 2 months CA. A specialist, Joëlle Provasi, blind to the infants’ group allocation and trained to the Expert level by the Einspieler team, analyzed the videos and calculated a Motor Optimality Score (MOS) for each infant to compare the mean of the MOS between the groups at term equivalent age and at 2 months CA.

### 6. Sample Size and Randomization

The planned 1600 home training sessions supervised by the 30 osteopaths constrained the sample size to 44. The stratified randomization process ensured we had a similar number of 24- 27+6 and 28-32 GA infants in each group.

### 7. Statistical methods

We used Statistica software (TIBCO Software Inc.) to run the analyses. Differences are given as mean differences with 95% confidence intervals and effect sizes are reported with confidence intervals when the effects are significant.

#### 7.1. Baseline data

We compared the perinatal characteristics of the infants and the parents’ education level among groups using separate univariate ANOVAs.

#### 7.2. Traveled distances during training

We compared the mean traveled distances by each infant for all the training sessions between the Crawli and Mattress groups using a student’s T-test and reported effect sizes using Cohen’s d.

#### 7.3. ATNAT and MOS scores

We compared the ATNAT and MOS scores among the three groups using separate univariate ANOVAs at the different ages tested because the total scores change according to the age at which the test is administered.

#### 7.4. Bayley Gross and Fine Motor Scaled Scores

##### 7.4.1. Primary analyses

The Bayley Gross and Fine Motor Scaled Scores at 2, 6, 9, and 12 months CA, which were normally distributed according to the Shapiro-Wilk statistic, were analyzed with 3(GROUP) x 4(AGE) ANOVAs with repeated measures on the AGE factor. We then performed separate one-way ANOVAs on GROUP at the four ages. Tukey’s tests were used to specify main effects. Differences are given as mean differences with 95% confidence intervals and effect sizes are reported using partial η^2^.

##### 7.4.2. Secondary Analyses

To deepen the Bayley Gross Motor results, we performed a second level of analysis of the Gross Motor scores by grouping certain items of interest reflecting the acquisition of a motor function: head holding, sitting, crawling, static standing, position transfers, and walking. For this purpose, the Gross Motor items grouped under the following functions are:

- *Head Control:* Item 3 “Lift his/her head less than 3 seconds from the shoulder of the experimenter who holds him/her vertically in his/her arms”; Item 4 “Lift his/her head for 3 seconds from the shoulder of the experimenter who holds him/her vertically in his/her arms”; Item 9 “Lift his/her head for 15 seconds from the shoulder of the experimenter who holds him/her vertically in his/her arms”; Item 11 “Stable head when carried vertically in the arms of an experimenter who is moving around the room”.
- *Sitting:* Item 22 “Sitting alone 5 seconds without support”; Item 26 “Sitting alone 30 seconds without support”; Item 27 “Sitting alone 60 seconds while manipulating an object”; Item 28 “Sitting alone while rotating the trunk to catch objects bilaterally”.
- *Crawling:* Item 30 “Belly-crawling for 1 meter”; Item 31 “Stand in a four-legged position”; Item 34 “Crawling on his or her hands and knees over 5 feet”.
- *Static Standing:* Item 33 “Standing for at least 2 seconds with light hand support”; Item 36 “Bouncing movements while standing with support of both hands”; Item 40 “Standing alone at least 3 seconds without support”.
- *Walking:* Item 37 “Walking with light hand support”; Item 42 “Three steps without support”; Item 43 “Walking at least five independent controlled steps”.
- *Transfers:* Item 41 “Lying on his/her back, the child is able to get up and stand, without any assistance or support. He/she first rolls to a prone or quadruped position and then stands up on his/her feet”; Item 46 “Lying on his/her back, the child is able to get up and stand, without any assistance or support and without rolling into the prone or quadruped position first”.

To perform the item grouping analysis for each stage of gross motor maturation, we summed the raw scores of the items comprising each grouping. Because the data were not distributed normally, we used a non-parametric Kruskal-Wallis test. We weighted ranks by the age of the subject (the youngest child having the highest rank). For this analysis, we compared the data from the Crawli group with those from the Control group and those from the Mattress group.

#### 7.5. Ages and Stages Questionnaires-3 (ASQ-3)

Consistent with our analysis of the Bayley Gross and Fine Motor scaled scores, we analyzed the Ages and Stages Questionnaires-3 scores at 9 and 12 months CA using 3(*GROUP*) x 2(*AGE*) ANOVAs with repeated measures on the *AGE* factor for the Total score and the scores for each of the five subscales of the questionnaire. We followed up any main effects of GROUP with Tukey tests. The Shapiro-Wilk test revealed the data were normally distributed. To provide greater specificity for the ASQ-3 results, we conducted separate one-way ANOVAs for the Total score and each of the subscale scores at 9 and 12 months CA. Tukey’s tests were used to specify main effects. Differences are given as mean differences with 95% confidence intervals and effect sizes are reported using partial η^2^.

## RESULTS

As some subjects missed some tests during the longitudinal follow-up, not all the participants were included in each analysis. However, we did not use imputation to replace missing data with substituted values. That accounts for the different degrees of freedom for the various tests.

### 1. Participant Flow and Recruitment

Among 107 participants assessed for eligibility, 49 infants were included and randomly assigned to the *Crawli* (n=17), *Mattress* (n=15) or *Control* (n=17) group. Five infants were excluded immediately after inclusion. Forty-four subjects were included in the final analysis with 15 in the *Crawli* group, 14 in the *Mattress* group and 15 in the *Control* group (See full description in Figure 4). The participants were predominantly Caucasian 11/15, 10/14 and 11/15 respectively in the Crawli, Mattress and Control groups. The others were of North or Central African descent.

**Figure 4.**
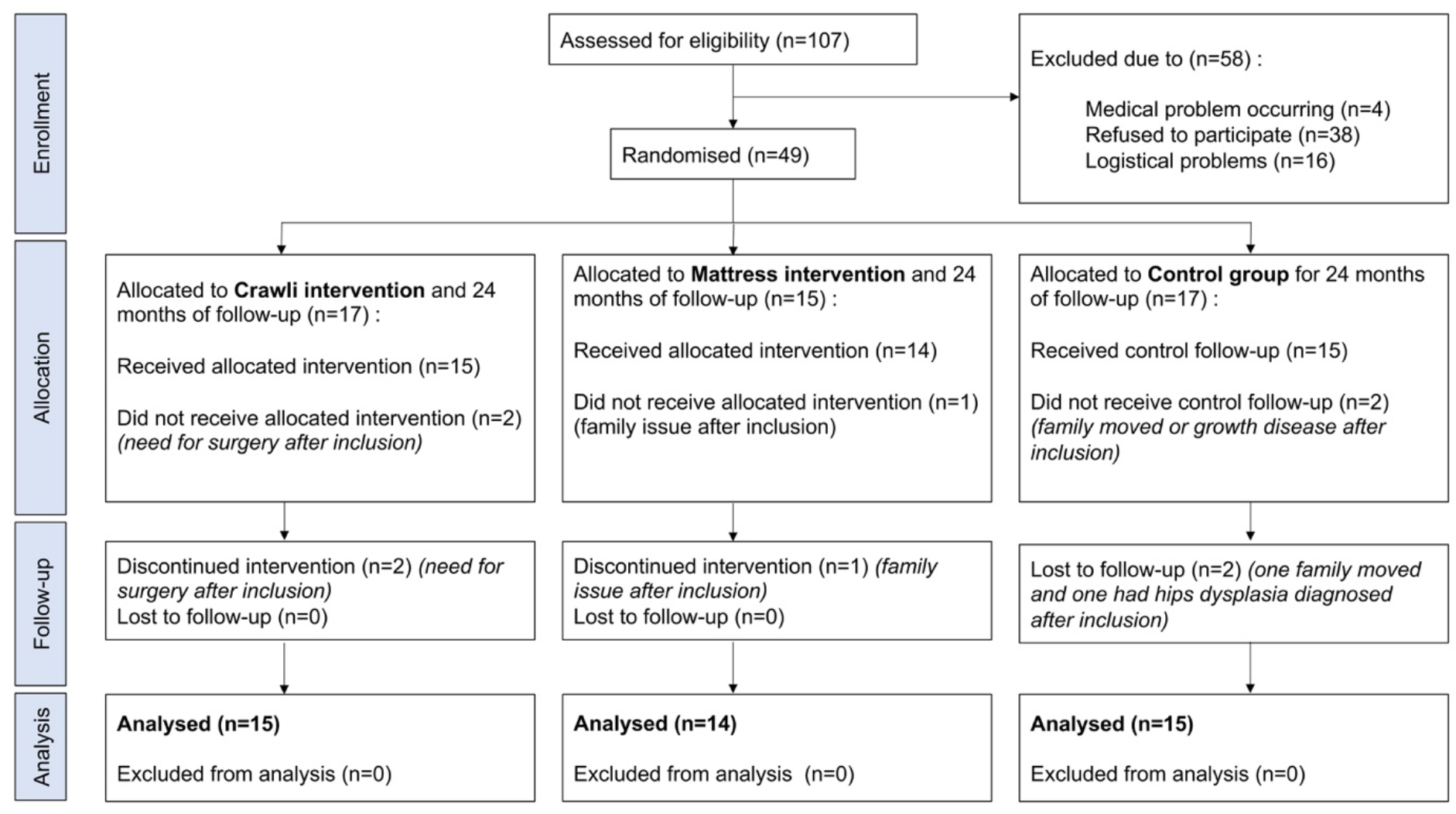
Flow chart CONSORT.

### 2. Baseline Data

There was no difference in perinatal characteristics and parental education level between groups (See full description in Table 1). There was no difference in either the ATNAT scores (F(2,41)=0.21, p=0.81, partial η^2^=0.01; Figure 5) or the Motor Optimality scores (F(2,33)=1.71, p=0.20, partial η^2^=0.93; Table 2) between groups at inclusion, confirming the homogeneity of the groups at inclusion.

**Figure 5.**
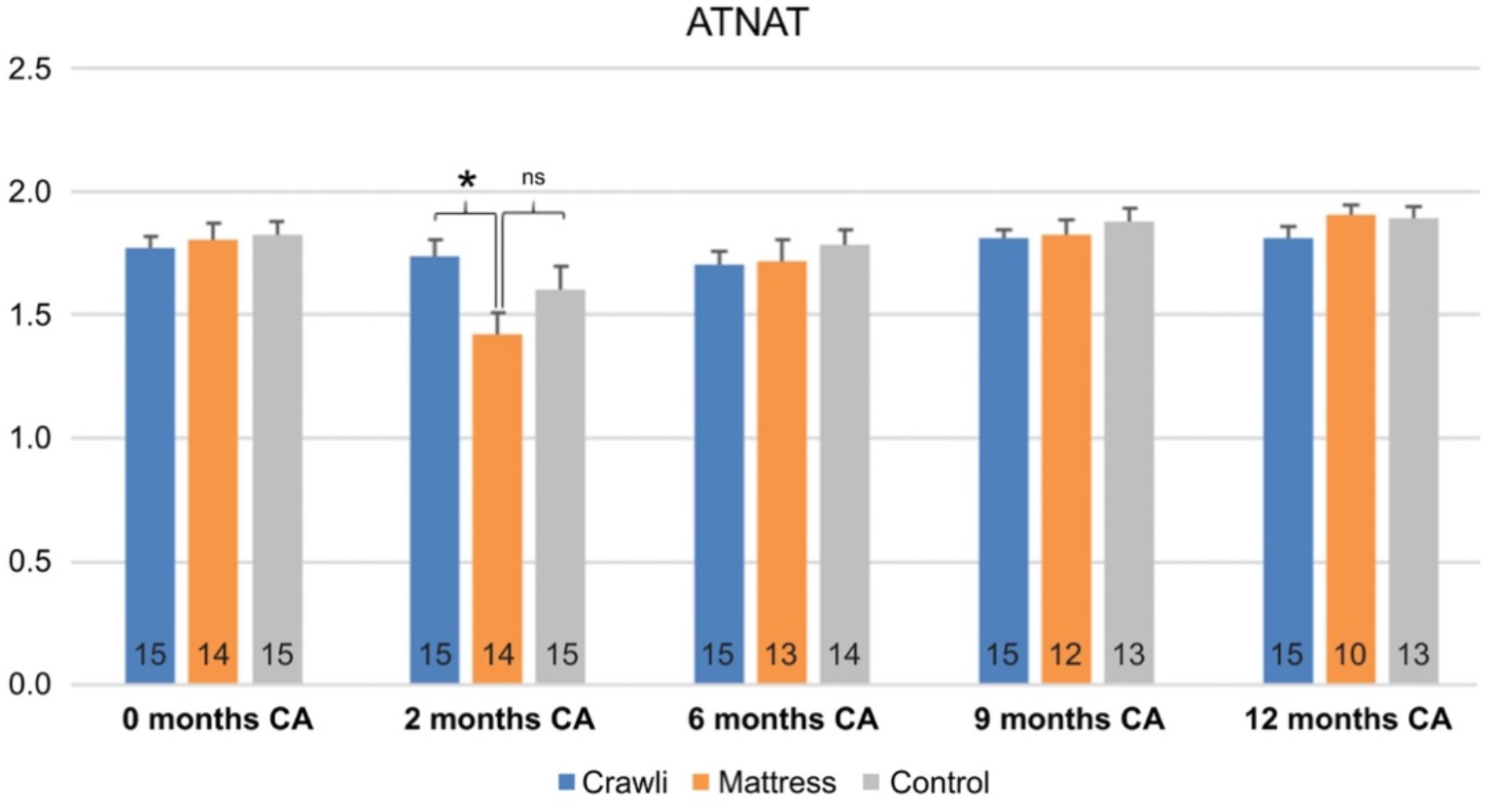
Comparison of ATNAT scores between groups at each age. The number of subjects is displayed at the bottom of each histogram. A score of 2 (on the y-axis) is considered as an optimal ATNAT score. Ages are in corrected age. Significant differences are indicated by * for p<.05 and ns indicates a non-significant difference.

**Table 1.** Morphological characteristics (GA gestational age, BW birthweight, HC head circumference) and parents level of education. In brackets we provide the percentages or standard deviations. Parental level of education is given in number of years of higher education. From row 1 downward, we obtained the following statistical results: 1) F (2,41) = 0.19, p = 0.83, partial η2 = 0.01; 2) F (2,41) = 0.25, p = 0.78, partial η2 = 0.01; 3) F (2,41) = 1.26, p = 0.29, partial η2 = 0.06; 4) F (2,41) = 0.29, p = 0.75, partial η2 = 0.01; 5) F (2,41) = 0.08, p = 0.92, partial η2 = 0.004; 6) F (2,41) = 0.34, p = 0.72, partial η2 = 0.02; 7) F (2,36) = 0.05, p = 0.95, partial η2 = 0.003; 8) F (2,41) = 0.88, p = 0.42, partial η2 = 0.04; 9) F (2,39) = 0.03, p = 0.97, partial η2 = 0.002; 10) F (2,39) = 2.42, p = 0.10, partial η2 = 0.11; 11) F (2,38) = 2.93, p = 0.07, partial η2 = 0.13; 12) F (2,39) = 2.1, p = 0.14, partial η2 = 0.10; 13) F (2,40) = 0.30, p = 0.75, partial η2 = 0.01; 14) F (2,37) = 0.97, p = 0.39, partial η2 = 0.05. Statistical differences are indicated by * (p<.05); and ns indicates a non- significant difference.

|  | Crawli Group | Mattress Group | Control Group | *P* |
| --- | --- | --- | --- | --- |
| **Number of subjects** | 15 | 14 | 15 |  |
| Subjects born < 28 weeks of GA | 3 (20%) | 4 (29%) | 3 (20%) | ns |
| Subjects BW<1000g | 4 (27%) | 3 (21%) | 5 (33%) | ns |
| Number of twins | 10 (67%) | 6 (43%) | 6 (40%) | ns |
| Proportions of boys | 6 (40%) | 5 (36%) | 4 (27%) | ns |
| **Morphological characteristics at birth** |  |  |  |  |
| Mean term age (GA+D) | 29+3 | 29+3 | 29+3 | ns |
| Mean birth weight (g) | 1202 (299.7) | 1294 (321.7) | 1227 (316.5) | ns |
| Mean birth height (cm) | 37.8 (3.6) | 37.6 (3.3) | 37.4 (3.2) | ns |
| Mean birth head circumference (cm) | 27.0 (2.4) | 27.7 (4.4) | 26.2 (1.8) | ns |
| **Morphological characteristics at inclusion** |  |  |  |  |
| Mean corrected age at inclusion (GA+D) | 39+5 | 39+6 | 39+6 | ns |
| Mean weight at inclusion (g) | 2686 (331.6) | 2680 (421.6) | 2436 (218.8) | ns |
| Mean height at inclusion (cm) | 44.5 (2.2) | 46.2 (1.7) | 44.4 (2.0) | ns |
| Mean head circumference at inclusion (cm) | 33.1 (0.8) | 33.3 (1.8) | 32.3 (1.3) | ns |
| **Parental level of education** |  |  |  |  |
| Educational levels of mothers | 4.5 (2.6) | 5.1 (3.5) | 4.3 (3.0) | ns |
| Educational levels of fathers | 5.2 (2.3) | 4.8 (2.3) | 3.8 (3.5) | ns |

**Table 2.** Motor Optimality Score (MOS) at term equivalent age (i.e., moment of inclusion in the study). Results of comparisons are presented as averaged differences between two groups. Statistics used were univariate analyses of variance. ns: non-significant. CA: corrected age.

| Motor Optimality Score (MOS) | Number of subjects | Mean (standard-deviation) | Confidence interval (95%) | Comparison with the mattress group | Comparison with the control group |
| --- | --- | --- | --- | --- | --- |
| **0 months CA** |  |  |  |  |  |
| Crawli group | 15 | 28.8 (8.7) | [24.0; 33.6] | [-7.0; 9.1]<sup>ns</sup> | [-14.7; 2.0]<sup>ns</sup> |
| Mattress group | 11 | 30.3 (10.9) | [23.0; 37.6] | - | [-16.3; 1.5]<sup>ns</sup> |
| Control group | 10 | 23.0 (11.0) | [15.1; 30.9] | - | - |

### 3. Training Adherence, Traveled distances during the sessions and Possible Harms

Adherence to the training was high, likely because the osteopaths were the ones who conducted the training in the infants’ homes each day and this motivated the parents’ compliance with the study. Of the 56 sessions, the Crawli group and Mattress groups completed an average of 52 (SD=2) and 51 (SD=2) sessions, respectively. The difference was not significant. The maximum number of sessions missed by a single infant was nine.

All infants trained in the Crawli group were able to move forward on the Crawliskate with a mean traveled distance per session of 138.7cm (SD=61.2) and a range from 68.1 to 242.3cm (see Figure 6A). As expected, in contrast to the Crawli group, infants positioned prone on the mattress were only able to move between 0.12 and 12.7 cm (mean=6.4cm, SD=4.4cm) (T(27)=8.07, p<0.00001, Cohen’s d = 3.0 (CI 95%[1.91- 4.06]); Figure 6A). We observed a high variability in the mean distances traveled by the infants on the Crawliskate during each of the eight weeks (Figure 6B).

**Figure 6.**
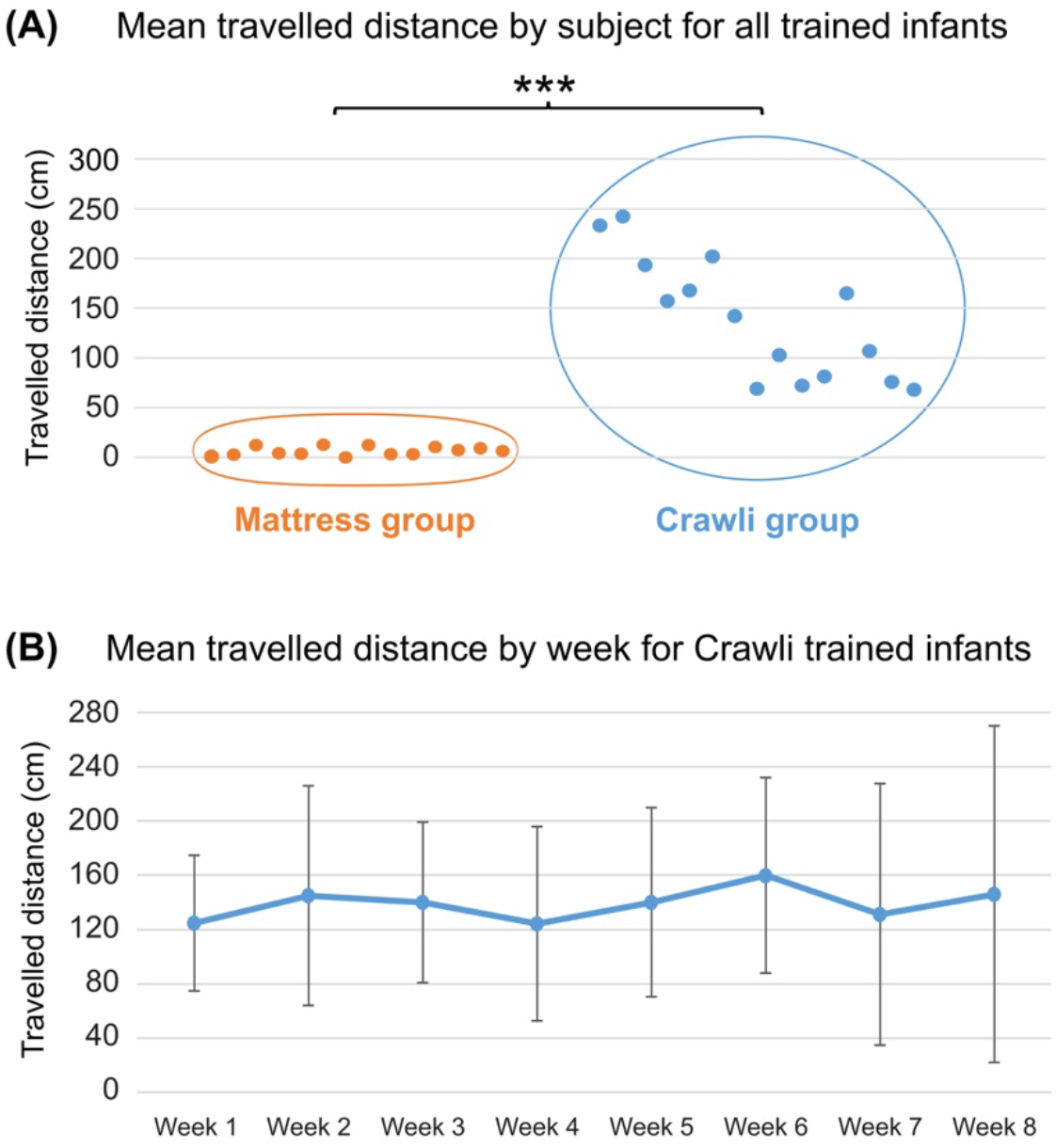
(A) Mean distance covered per session by each subject in each group (Orange = Mattress group; Blue = Crawli group) during his/her entire training. The distance covered (in cm) is represented on the y-axis and the subjects are distributed on the x-axis. The *** indicates a p-value <0.001. **(B)** Graphical representation of the mean distance (and SD) covered during each of the eight weeks of training for the Crawli group.

The osteopaths reported no harms to the trained infants and the ATNAT follow-up assessment at each age up to 12 months CA showed no deleterious effects of training in the Crawli and Mattress groups: similar ATNAT scores were observed between Crawli, Mattress and Control groups except for an unexpected lower score for the Mattress group than the Crawli group at 2 months CA (F(2,40)=3.62, p=0.04, partial η2=0.15 (CI 95%[0.08-0.55]); HSD Tukey Crawli > Mattress, p=0.03; Mattress=control, p=0.28 and Crawli=control, p = 0.51) but not thereafter (ATNAT 6: F(2,40)=.13, p=0.88, partial η2=0.01; ATNAT 9: F(2,39)=1.00, p=0.37, partial η2=0.05; ATNAT 12: F(2,38)=0.98, p=0.39, partial η2=0.05; Figure 5). The temporary lower score for the Mattress group at 2 months CA was due to a greater spinal axis hyper extension on the corresponding specific ATNAT item on spinal axis (data not shown). The groups demonstrated similar MOS at 2 months CA (F (2,38)=1.32, p=0.28, partial η2=0.06; Table 3).

**Table 3.**
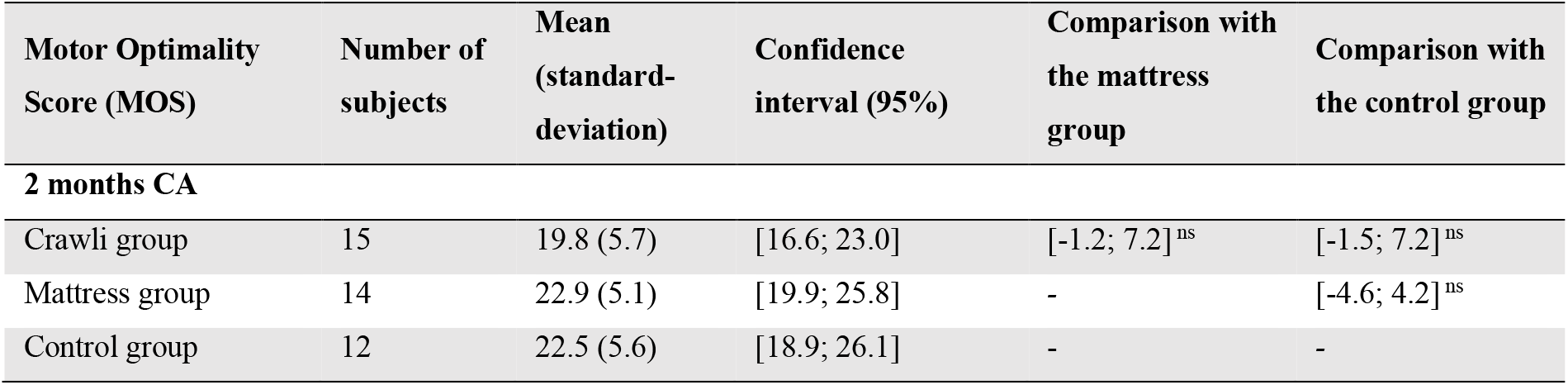
Motor Optimality Score (MOS) at 2 months corrected age (CA). Results of comparisons are presented as averaged differences between two groups. ns: non-significant.

| Motor Optimality Score (MOS) | Number of subjects | Mean (standard-deviation) | Confidence interval (95%) | Comparison with the mattress group | Comparison with the control group |
| --- | --- | --- | --- | --- | --- |
| **2 months CA** |  |  |  |  |  |
| Crawli group | 15 | 19.8 (5.7) | [16.6; 23.0] | [-1.2; 7.2]<sup>ns</sup> | [-1.5; 7.2]<sup>ns</sup> |
| Mattress group | 14 | 22.9 (5.1) | [19.9; 25.8] | - | [-4.6; 4.2]<sup>ns</sup> |
| Control group | 12 | 22.5 (5.6) | [18.9; 26.1] | - | - |

### 4. Bayley Gross Motor Scaled Scores

All Gross Motor (GM) scaled scores are summarized in Figure 7 and Table 4.

**Figure 7.**
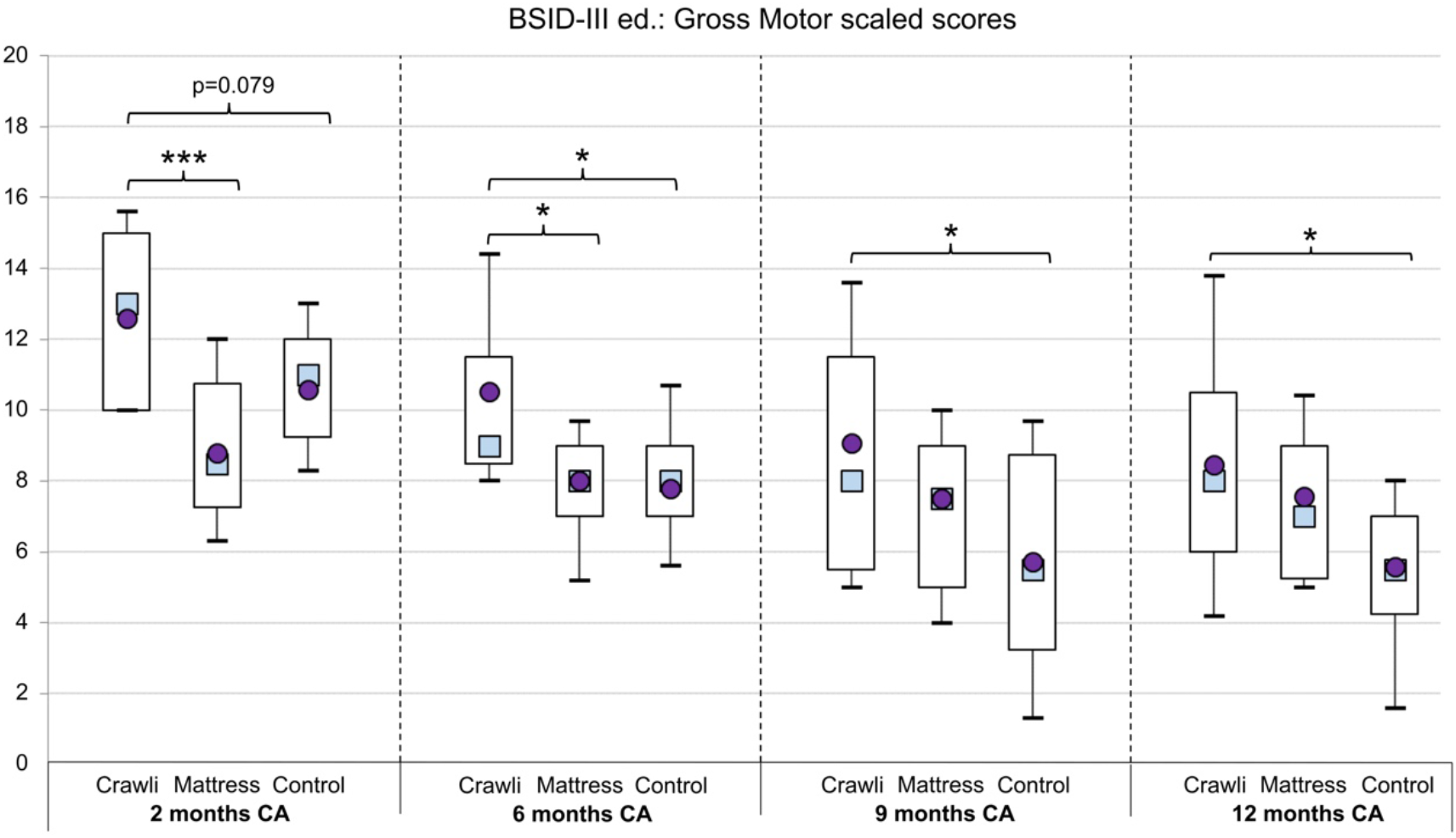
Gross motor scaled score. The whisker boxes represent the 10th, 25th, 75th and 90th percentiles with the mean represented by a circle and the median by a square. Significant differences are indicated with * for p<0.05; ** for p<0.01; *** for p<0.001; p-values between 0.05 and 0.1 are reported in superscript.

**Table 4.** Bayley gross motor (GM) scaled score (SS) at corrected age (CA). Mean diff. represents mean difference of scoring between two groups, statistical analysis was conducted with one-way ANOVAs. Significant differences and trends are in bold: * for p<0.05; ** for p<0.01; *** for p<0.001; A p-value between 0.05 and 0.1 is reported in superscript.

| Bayley Gross Motor Scaled Score | Number of Subjects | Mean (standard deviation) | Confidence intervals (95%) | Mean diff. with Mattress | Mean diff. with Control |
| --- | --- | --- | --- | --- | --- |
| **At 2 months CA** |  |  |  |  |  |
| Crawli group | 15 | 12.6 (2.7) | [11.1; 14.1] | **[1.91; 5.72]\*\*\*** | **[0.20; 4.27]<sup>0.08</sup>** |
| Mattress group | 14 | 8.8 (2.4) | [7.4; 10.2] | - | [-0.49; 3.64] |
| Control group | 14 | 10.6 (2.2) | [9.3; 11.9] | - | - |
| **At 6 months CA** |  |  |  |  |  |
| Crawli group | 15 | 10.5 (3.2) | [8.8; 12.3] | **[0.62; 4.44]\*** | **[0.85; 4.94]\*** |
| Mattress group | 14 | 8.0 (1.7) | [7.1; 8.9] | - | [-2.43; 1.71] |
| Control group | 14 | 7.8 (2.5) | [6.3; 9.3] | - | - |
| **At 9 months CA** |  |  |  |  |  |
| Crawli group | 15 | 9.1 (3.8) | [6.9; 11.2] | [-0.88; 4.01] | **[1.55; 6.77]\*** |
| Mattress group | 14 | 7.5 (2.6) | [6.0; 9.0] | - | [-5.24; 0.06] |
| Control group | 14 | 5.7 (3.3) | [3.8; 7.6] | - | - |
| **At 12 months CA** |  |  |  |  |  |
| Crawli group | 15 | 8.5 (3.8) | [6.3; 10.6] | [-1.49; 3.28] | **[0.55; 5.65]\*** |
| Mattress group | 14 | 7.6 (2.2) | [6.3; 8.8] | - | [-4.79; 0.38] |
| Control group | 14 | 5.6 (2.9) | [3.9; 7.2] | - | - |

#### 4.1. Comprehensive ANOVA

The main effect of *GROUP* was significant, (F(2,37)=7.39, p<0.001), partial η^2^=0.28 (CI 95% [0.05-0.46]). Tukey’s test revealed the Crawli group scaled score was significantly higher than that of the Control group (p<0.01) and the Mattress group (p<0.05). The difference between the Control and Mattress group was not significant (p=0.6). The main effect of AGE was significant, (F(3,111)=21.07, p<0.0001), partial η^2^=0.36 (CI 95% [0.21-0.47]), and the *AGE x GROUP* interaction was significant, (F(6,111)=2.53, p=0.02), partial η^2^=0.12 (CI 95% [0-0.20]). However, because we used the scaled Bayley Gross Motor score, we did not use the Tukey test to follow up on the main effect of *AGE* in the ANOVA. Comparison of scaled scores across age groups is not particularly meaningful.

#### 4.2. Univariate ANOVA at 2 months CA

The ANOVA at 2 months of corrected age (CA) revealed a significant *GROUP* effect (F(2,40)=8.68, p<0.001), partial η^2^=0.30 (CI 95% [0.07-0.47]). The post-hoc test revealed the Crawli group scored significantly higher than the Mattress group (p<0.001) and there was a trend for the Crawli group to score higher than the Control group (p=0.08).

#### 4.3. Univariate ANOVA at 6 months CA

The ANOVA at 6 months CA revealed a significant *GROUP* effect (F(2,40)=5.30, p<0.01), partial η^2^=0.21 (CI 95% [0.02-0.39]). The post-hoc test revealed the Crawli group scores were significantly higher than those of the Control group (p<0.05) and the Mattress group (p<0.05).

#### 4.4. Univariate ANOVA at 9 months CA

The ANOVA at 9 months CA revealed a significant *GROUP* effect (F(2,40)=3.74, p<0.05), partial η^2^=0.16 (CI 95% [0-0.33]). The post-hoc test revealed the Crawli group scores were significantly higher than those of the Control group (p<0.05).

#### 4.5. Univariate ANOVA at 12 months CA

The ANOVA at 12 months CA revealed a significant *GROUP* effect (F(2,40)=3.33, p<0.05), partial η^2^=0.14 (CI 95% [0-0.32]). The post-hoc test revealed the Crawli group scores were significantly higher than those of the Control group (p<0.05).

#### 4.6. Specific items

With reference to the secondary analyses of the Bayley items (Figure 8), the analyses revealed that the scores of the Crawli group were significantly higher 1) for head control at 2 months CA compared to the Mattress group (p<0.001; η^2^=0.62) and the Control group (p<0.05; η^2^=0.18), 2) for mature crawling at 9 months CA compared to the Mattress group (p<0.05; η^2^=0.11) and the Control group (p<0.05; η^2^=0.12), 3) for static standing (p<0.01; η^2^=0.23) and the position transfers (p<0.01; η^2^=0.26) compared to the Control group at 12 months CA. The Crawli group had scores higher than the Mattress group for sitting at 6 months CA (tendency p=0.066; η^2^=0.09) and higher than the Control group for walking at 9 months CA (tendency p=0.066; η^2^=0.09).

**Figure 8.**
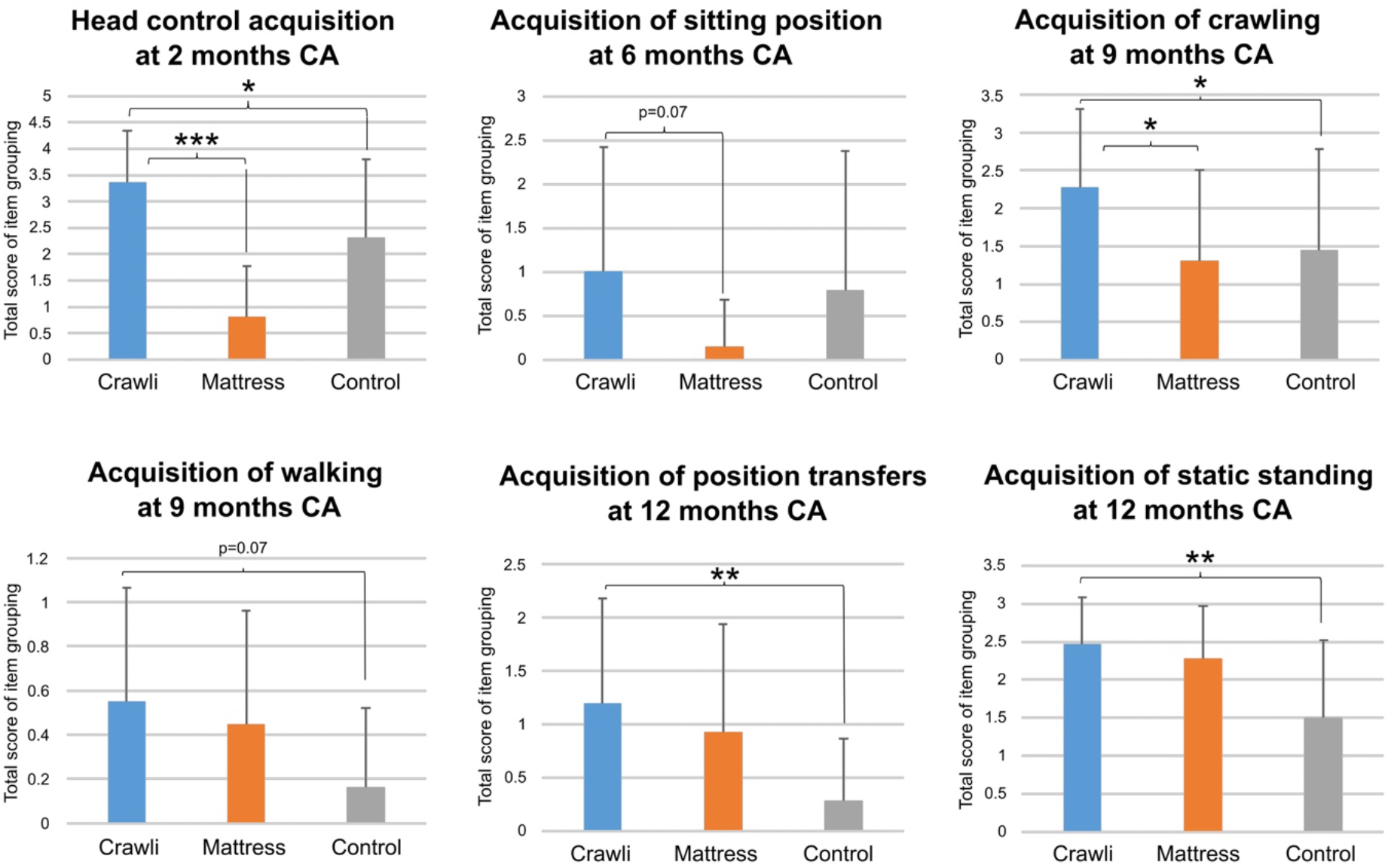
Graphical representation of raw scores of grouped items of the Bayley scales reflecting major gross motor skills. Statistical analyses were done using Kruskal-Wallis tests. Significant differences and trends are shown in bold: * for p <0.05; ** for p <0.01; *** for p <0.001.

### 5. Bayley Fine Motor Scaled Scores

The 3 groups were similar in their Bayley Fine Motor scaled scores (results are summarized in Table 5).

**Table 5.** Bayley Fine Motor (FM) Scaled Scores (SS) at corrected age (CA). Mean diff. represents mean difference of scoring between two groups, statistical analysis was conducted with one-way ANOVAs. ns: non-significant.

| **BSID Fine Motor Scaled Score** | **Number of subjects** | **Mean (standard-deviation)** | **Confidence interval (95%)** | **Mean diff. with Mattress** | **Mean diff. with Control** |
| --- | --- | --- | --- | --- | --- |
| **2 months CA** |  |  |  |  |  |
| Crawli group | 15 | 13.3 (2.0) | [12.2; 14.4] | [-3.8; 0.07]<sup>ns</sup> | [-2.4; 2.0]<sup>ns</sup> |
| Mattress group | 14 | 11.1 (3.2) | [9.3; 13.0] | - | [-0.7; 4.1]<sup>ns</sup> |
| Control group | 11 | 12.4 (2.4) | [10.8; 14.0] | - | - |
| **6 months CA** |  |  |  |  |  |
| Crawli group | 15 | 10.9 (2.9) | [9.3; 12.5] | [-4.5; 0.0]<sup>ns</sup> | [-4.0; 1.2]<sup>ns</sup> |
| Mattress group | 13 | 8.0 (3.9) | [5.7; 10.3] | - | [-1.9; 3.6]<sup>ns</sup> |
| Control group | 14 | 8.9 (2.3) | [7.3 ; 10.6] | - | - |
| **9 months CA** |  |  |  |  |  |
| Crawli group | 15 | 11.3 (2.5) | [9.9; 12.7] | [-2.7; 1.1]<sup>ns</sup> | [-3.3; 1.0]<sup>ns</sup> |
| Mattress group | 12 | 10.2 (1.9) | [9.0; 11.3] | - | [-2.7; 1.9]<sup>ns</sup> |
| Control group | 14 | 10.7 (2.9) | [9.1; 12.4] | - | - |
| **12 months CA** |  |  |  |  |  |
| Crawli group | 15 | 10.3 (3.3) | [8.4; 12.1] | [-2.4; 2.4]<sup>ns</sup> | [-1.6; 3.9]<sup>ns</sup> |
| Mattress group | 11 | 10.3 (3.0) | [8.3; 12.3] | - | [-1.8; 4.1]<sup>ns</sup> |
| Control group | 13 | 12.4 (2.7) | [10.8; 14.0] | - | - |

#### 5.1. Comprehensive ANOVA

The main effect of *GROUP* was not significant, (F(2,37)=1.07, p=0.38, partial η2=0.07). The main effect of AGE was significant, (F(3,90)=6.06, p=0.0008, partial η2=0.17 (CI 95% [0.03-0.28]) and the *AGE x GROUP* interaction was not significant, (F(2,37)=0.94, p=0.45 partial η2=0.06). However, because we used the scaled Bayley Fine Motor score, we did not use the Tukey test to follow up on the main effect of *AGE* in the ANOVA. Comparison of scaled scores across age groups is not particularly meaningful. Despite the non-significant findings for GROUP and the AGE x GROUP interaction, we conducted the univariate ANOVAs at each age group for the sake of completeness.

#### 5.2. Univariate ANOVA at 2 months CA

The ANOVA at 2 months of corrected age (CA) did not reveal a significant *GROUP* effect (F(2,37)=2.62, p=0.09, partial η^2^ = 0.12).

#### 5.3. Univariate ANOVA at 6 months CA

The ANOVA at 6 months CA did not reveal a significant *GROUP* effect (F(2,39)=3.08, p=0.06, partial η^2^=0.14).

#### 5.4. Univariate ANOVA at 9 months CA

The ANOVA at 9 months CA did not reveal a significant *GROUP* effect (F(2,38)=0.75, p=0.48, partial η2=0.04).

#### 5.5. Univariate ANOVA at 12 months CA

The ANOVA at 12 months CA did not reveal a significant *GROUP* effect (F(2,36)=2.10, p=0.14, partial η2=0.10).

### 6. ASQ-3 Scores

ASQ-3 results are summarized in Tables 6 and 7.

**Table 6.** Ages and Stages Questionnaire – 3. Results at 9 months corrected age. Mean diff. represents mean difference of scoring between two groups, statistical analyses were conducted with one-way ANOVAs. Significant differences and trends are in bold; * for p<0.05; ** for p<0.01; *** for p<0.001. A p-value between 0.05 and 0.1 is reported in superscript.

| Ages & Stages Questionnaire - 3<br>At 9 months corrected age |  | Mean (standard<br>deviation) | Confidence<br>Intervals (95%) | Mean diff.<br>with Mattress | Mean diff.<br>with Control |
| --- | --- | --- | --- | --- | --- |
| **TOTAL SCORE** | Crawli (n=13) | 225.38 (53.05) | [193.33; 257.44] | [-1.51; 68.95] | **[23.94; 92.98]\*\*** |
|  | Mattress (n=12) | 191.67 (37.31) | [167.96; 215.38] | - | [-10.48; 59.97] |
|  | Control (n=13) | 166.92 (37.28) | [144.39; 189.45] | - | - |
| **Gross motor** | Crawli | 38.46 (18.97) | [27.00; 49.93] | [-5.88; 21.14] | **[4.07; 30.54]\*** |
|  | Mattress | 30.83 (14.43) | [21.66; 40.00] | - | [-3.83; 23.19] |
|  | Control | 21.15 (15.96) | [11.51; 30.80] | - | - |
| **Fine motor** | Crawli | 55.77 (5.34) | [52.54; 59.00] | [0.28; 20.42] | **[2.06; 21.79]\*** |
|  | Mattress | 45.42 (9.40) | [39.44; 51.39] | - | [-8.50; 11.64] |
|  | Control | 43.85 (18.39) | [32.73; 54.96] | - | - |
| **Problem solving** | Crawli | 43.08 (15.75) | [33.56; 52.60] | [-14.15; 8.63] | [-6.93; 15.39] |
|  | Mattress | 45.83 (7.64) | [40.98; 50.69] | - | [-4.40; 18.38] |
|  | Control | 38.85 (16.48) | [28.89; 48.80] | - | - |
| **Communication** | Crawli | 48.46 (13.13) | [40.53; 56.40] | **[0.85; 20.24]<sup>0.08</sup>** | **[4.73; 23.73]\*** |
|  | Mattress | 37.92 (11.96) | [30.32; 45.51] | - | [-6.01; 13.38] |
|  | Control | 34.23 (10.58) | [27.84; 40.62] | - | - |
| **Personal social** | Crawli | 39.61 (12.82) | [31.87; 47.36] | [-1.81; 17.71] | **[1.21; 20.33]<sup>0.07</sup>** |
|  | Mattress | 31.67 (11.55) | [24.33; 39.00] | - | [-6.94; 12.58] |
|  | Control | 28.85 (11.57) | [21.85; 35.84] | - | - |

**Table 7.** Ages and Stages Questionnaire – 3. Results at 12 months corrected age. Mean diff. represents mean difference of scoring between two groups, statistical analyses were conducted with one-way ANOVAs. Significant differences and trends are in bold; * for p<0.05; ** for p<0.01; *** for p<0.001. A p-value between 0.05 and 0.1 is reported in superscript.

| Ages & Stages Questionnaire - 3<br>At 12 months corrected age |  | Mean (standard<br>deviation) | Confidence<br>Intervals (95%) | Mean diff.<br>with Mattress | Mean diff.<br>with Control |
| --- | --- | --- | --- | --- | --- |
| **TOTAL SCORE** | Crawli (n=15) | 240.33 (32.92) | [222.10; 258.56] | [-1.32; 58.42] | **[15.10; 74.85]\*** |
|  | Mattress (n=14) | 211.79 (41.07) | [188.07; 235.50] | - | [-13.95; 46.81] |
|  | Control (n=14) | 195.36 (44.87) | [169.45; 221.26] | - | - |
| **Gross motor** | Crawli | 41.67 (17.29) | [32.09; 51.24] | [-8.21; 20.83] | [-0.35; 28.68] |
|  | Mattress | 35.36 (20.98) | [23.24; 47.47] | - | [-6.91; 22.63] |
|  | Control | 27.50 (19.68) | [16.13; 38.87] | - | - |
| **Fine motor** | Crawli | 56.33 (5.50) | [53.29; 59.38] | **[2.28; 17.52]\*** | **[3.36; 18.62]\*** |
|  | Mattress | 46.43 (9.29) | [41.07; 51.79] | - | [-6.67; 8.82] |
|  | Control | 45.36 (14.07) | [37.23; 53.48] | - | - |
| **Problem solving** | Crawli | 50.57 (7.76) | [46.37; 54.96] | **[1.28; 17.20]<sup>0.06</sup>** | [-0.51; 15.41] |
|  | Mattress | 41.43 (7.70) | [36.98; 45.88] | - | [-9.88; 6.31] |
|  | Control | 43.21 (14.89) | [34.62; 51.81] | - | - |
| **Communication** | Crawli | 46.67 (10.46) | [40.87; 52.46] | [-8.22; 7.98] | **[1.07; 17.26]<sup>0.07</sup>** |
|  | Mattress | 46.79 (11.37) | [40.22; 53.35] | - | **[1.05; 17.52]<sup>0.07</sup>** |
|  | Control | 37.50 (10.52) | [31.43; 43.57] | - | - |
| **Personal social** | Crawli | 45.00 (8.24) | [40.44; 49.56] | [-4.72; 11.15] | [-4.72; 11.15] |
|  | Mattress | 41.79 (10.12) | [35.95; 47.63] | - | [-8.07; 8.07] |
|  | Control | 41.79 (12.96) | [34.31; 49.26] | - | - |

### 6.1. Comprehensive ANOVA for Total score

The main effect for *GROUP* (F(2,35)=6.02, p<0.01), partial η^2^=0.26 (CI 95% [0.03-0.44]), was significant for the Total score. The post-hoc test revealed the Crawli total score was significantly higher than that of the Control group (p<0.01). The difference between the Crawli and Mattress group and between the Control and Mattress group were not significant (respectively p=0.1 and p=0.4). The main effect for *AGE* (F(1,35)=15.81, p<0.001), partial η^2^=0.31 (CI 95% [0.08-0.50]) was also significant but not the *AGE x GROUP* interaction (F(2,35)=0.35, p=0.71), partial η^2^=0.02). However, because the Ages and Stages Questionnaires scores are also scaled according the child’s age, we did not use the Tukey test to follow up on the main effect of *AGE* in the ANOVA. Comparison of scores across age groups is not particularly meaningful.

### 6.2. Comprehensive ANOVA for Gross Motor Subscale

For the Gross Motor subscale, neither the main effect for *GROUP* (F(2,35)=2.75, p=0.08; partial η^2^=0.14), nor *AGE* (F(1,35)=3.06, p=0.09, partial η^2^=0.08), nor the *AGE x GROUP* interaction were significant (F(2,35)=0.41, p=0.67, partial η^2^=0.02).

### 6.3. Comprehensive ANOVA for Fine Motor Subscale

For the Fine Motor subscale, the main effect of *GROUP* was significant, (F(2,35)=6.75, p<0.01), partial η^2^=0.28 (CI 95% [0.40-0.46]), however, neither the main effect of *AGE* (F(1,35)=0.45, p=0.51, partial η^2^=0.01), nor the *AGE x GROUP* interaction were significant (F(2,35)=0.03, p=0.97), partial η^2^=0.001). The post-hoc test revealed the Crawli group scored significantly higher than the Mattress group (p<0.05) and the Control group (p<0.01).

### 6.4. Comprehensive ANOVA for Problem Solving Subscale

For the Problem Solving subscale, neither the main effect of *GROUP* (F(2,35)=1.28, p=0.29, partial η^2^=0.07), nor *AGE* (F(1,35)=0.82, p=0.37, partial η^2^=0.02), nor the *AGE x GROUP* interaction (F(2,35)=2.43, p=0.10; partial η^2^=0.12) were significant.

### 6.5. Comprehensive ANOVA for Communication Subscale

For the Communication subscale, the main effect of *GROUP* (F(2,35)=4.05, p<0.05), partial η^2^=0.19 (CI 95% [0-0.37]) and *AGE* (F(1,35)=5.78, p<0.05), partial η^2^=0.14 (CI 95% [0-0.35]) and the *AGE x GROUP* interaction (F(2,35)=3.54, p<0.05), partial η^2^=0.17 (CI 95% [0-0.35]) were significant. The post-hoc test revealed the Crawli group scored significantly higher than the Control group (p<0.05).

### 6.6. Comprehensive ANOVA for Personal Social Subscale

For the Personal Social subscale, the main effect of *AGE* (F(1,35)=20.38, p<0.001), partial η^2^=0.37 (CI 95% [0.12- 0.55]) was significant, however neither the main effect of *GROUP* (F(2,35)=1.99, p=0.15, partial η^2^=0.10) nor the *AGE x GROUP* interaction (F(2,35)=1.02, p=0.37, partial η^2^=0.05) were significant.

### 6.7. Univariate ANOVAs at 9 months CA

For the *Total score* at 9 months CA, the ANOVA was significant (F(2,35)=5.95, p<0.01), partial η^2^=0.25 (CI 95% [0.03-0.44]). The post-hoc test revealed the Crawli group scored significantly higher than the Control group (p<0.01). For the *Gross Motor score* at 9 months CA, the ANOVA was significant (F(2,35)=3.54, p<0.05), partial η^2^=0.17 (CI 95% [0-0.35]). The post-hoc test revealed the Crawli group scored significantly higher than the Control group (p<0.05). For the *Fine Motor score* at 9 months CA, the ANOVA was significant (F(2,35)=3.52, p<0.05), partial η^2^=0.17 (CI 95% [0-0.35]). The post-hoc test revealed the Crawli group scored significantly higher than the Control group (p<0.05). For the *Problem Solving score* at 9 months CA, the ANOVA was not significant (F(2,35)=0.79, p=0.46, partial η^2^=0.04). For the *Communication score* at 9 months CA, the ANOVA was significant (F(2,35)=4.96, p<0.05), partial η^2^=0.22 (CI 95% [0.01-0.41]). The post-hoc test revealed the Crawli group scored significantly higher than the Control group (p<0.05). There was a trend for the Crawli group to score higher than the Mattress group (p=0.08). For the *Personal Social score* at 9 months CA, the ANOVA trended toward significance (F(2,35)=2.80, p=0.07), partial η^2^=0.14 (CI 95% [0-0.32]). The Crawli group scores were almost significantly higher than those of the Control Group (p=0.07).

### 6.8. Univariate ANOVAs at 12 months CA

For the *Total score* at 12 months CA, the ANOVA was significant (F(2,40)=4.77, p<0.05), partial η^2^=0.19 (CI 95% [0.01-0.37]). The post-hoc test revealed the Crawli group scored significantly higher than the Control group (p<0.05). For the *Gross Motor score* at 12 months CA, the ANOVA was not significant (F(2,40)=1.95, p=0.16, partial η^2^=0.09). For the *Fine Motor score* at 12 months CA, the ANOVA was significant (F(2,40)=5.21, p<0.001), partial η^2^=0.21 (CI 95% [0.01-0.38]). The post-hoc test revealed the Crawli group scored significantly higher than the Control group (p<0.05) and the Mattress group (p<0.05). For the *Problem Solving score* at 12 months CA, the ANOVA trended towards significance (F(2,40)=3.13, p=0.05), partial η^2^=0.13 (CI 95% [0- 0.31]). The Crawli group scores tended to be higher than those of the Mattress group (p=0.06). For the *Communication score* at 12 months CA, the ANOVA was significant (F(2,40)=3.45, p<0.05), partial η^2^=0.15 (CI 95% [0-0.32]). However, the post-hoc test failed to identify the significant differences with the Crawli group scores tending to be higher than those of the Control group (p=0.07) and the Mattress group (p=0.07); and the Mattress group scores tending to be higher than those of the Control group (p=0.07). For the *Personal Social score* at 12 months CA, the ANOVA was not significant (F(2,40)=0.45, p=0.64, partial η^2^=0.02).

## DISCUSSION

Prematurity continues to increase at a constant rate, even though it appears to have stabilized recently in some countries (2). However, methods to train the locomotor and gross motor skills of premature infants at risk of disability at a very early age are sorely lacking (29). The goal of the present study was to test the feasibility of using a mini-skateboard, the Crawliskate, to train very premature infants without major brain injuries, but who were nevertheless at heightened risk for developmental delay and/or disability, to crawl daily in the home from term equivalent age onward and to determine the effects of this type of training on their motor and general development up to 12 months CA. Although we had a small sample size, our results show not only the feasibility of our intervention but more importantly that daily stimulation of the quadrupedal gait of premature infants upon discharge from the NICU has positive effects on their development of mature crawling at 9 months CA and on their gross motor and general development.

### Training adherence and possible harms

First, we observed excellent adherence to the training, likely because the osteopaths we recruited conducted the training in the infants’ homes each day. The results show the feasibility of such early home-based crawling training performed 5 min/day for 8 consecutive weeks following discharge from the NICU. Specifically, all infants in the Crawli groups were able to propel themselves using quadrupedal movements with the help of the Crawliskate without deleterious effects, as shown by similar scores on the ATNAT at 2, 6, 9 and 12 months CA and GMA at 2 months CA. As predicted, infants positioned directly on the mattress without the Crawliskate were unable to move very far and unexpectedly developed a temporary head and trunk hyper extension at 2 months CA.

### Traveled distances during Crawli training

It is remarkable that even at this very early age, premature infants could travel long distances with the help of the Crawliskate, up to a maximum of 2.5 meters in only 5 min for some of the infants. However, we observed a large variability within and between infants in traveled distance. We can likely attribute the variability in distance traveled to variation in the infant’s state of alertness at each of the training sessions. Sometimes infants appeared too tired or sleepy to move much during the session. We also saw variability within sessions, with infants often starting to crawl, then stopping, then resuming again at the end of the session. Consequently, we found it difficult to construct a learning profile for crawling during the 8 consecutive weeks of training for each infant. We are presently working to code the number of leg and arm steps during each session to determine the distance traveled per step, which should provide a better representation of the progression of crawling skill during the training.

### Effect of the neonatal crawling intervention on mature Crawling

One of the remarkable effects we saw from stimulating the very premature infants crawling at an early age was the impact it had on the development of mature crawling at 9 months CA, with infants in the Crawli group showing significantly more mature crawling than infants in the Mattress and Control groups. This result is not simply the consequence of prone positioning because the Mattress group infants demonstrated a similar level of crawling competence as infants in the Control group. The effect of early crawling training on mature crawling confirms, with a new paradigm, the existence of a link between neonatal and mature locomotion. Researchers have reported previously a link between stepping and walking by showing that daily stepping on a solid surface from birth enhances the emergence and quality of mature walking in typically-developing infants (57,58) and training stepping on a treadmill leads to an earlier emergence and higher quality of walking in infants with Down syndrome (59,60,62,63,77). Our results on crawling training also confirm, but with a much earlier intervention, results reported 40 years ago showing that daily training of supported crawling in typically developing infants from four months of age onward leads to an earlier emergence of independent crawling and walking (19). However, we did not find an effect of our intervention on walking, as scored by Bayley items at 12 months CA, only a strong trend for the Crawli infants to have higher scores relative to the Control group on the items related to walking at 9 months CA (p=0.07). This result for walking could result from the fact that, in contrast to typically-developing infants, very premature infants may need more than 5 minutes per day and/or more than 8 weeks of crawling training to accelerate the development of their walking proficiency. Alternately, the Bayley items related to walking at 12 months CA may lack the sensitivity to detect subtle differences in the quality of walking between groups of very preterm infants without brain damage. As we also collected the spatio-temporal parameters of these infants walking patterns on a GAITRite pressure sensitive walkway at 24 months CA, we may still find an effect of our intervention on walking.

### Effect of the Crawling Intervention on Gross Motor Development

The crawling intervention had positive effects on infants’ gross motor development when the Bayley scores were aggregated over the four age periods. When looking by age, the Crawli group had higher scores than the Control group at 6, 9 and 12 months CA, that is 10 months after the end of the crawling intervention. The Crawli group had higher scores than the Mattress group at 2 and 6 months CA. The similar gross motor scores between the Crawli and Mattress groups at 9 and 12 months CA may be the result of the limited duration of the training, although it is important to note (see the previous section) that the Crawli group had significantly higher scores on the specific Bayley items related to mature crawling at 9 months CA relative to the Mattress group. If the training lasted longer than two months, the group differences at 9 and 12 months may have been larger and more likely to be significant, particularly with a larger sample. However, we think a more likely explanation for the attenuated differences between Crawli and Mattress groups at older ages is the additional time parents in the Mattress group reported continuing to place their infants in the prone position for more than two hours per day after the training (58% and 60% for the Mattress group between 2 to 6 months CA and 6 to 9 months CA, respectively and 21% and 38% for the Crawli group during the same age periods). Tummy time, practiced when the infant is older, is known to have beneficial effects on infant motor development (70). Although the Bayley Gross Motor and ASQ-3 scores of the Crawli and Mattress groups did not differ significantly between 9 and 12 months CA, it is pertinent to note that the Crawli group’s scores were always higher than the Mattress group’s scores. Moreover, the Mattress group’s scores were never significantly higher than the Control group’s scores at any age tested.

The generalized and enduring effects of the crawling training on the infants’ motor development, particularly relative to the Control group, are quite surprising in the context of prior work which generally shows that motor practice effects are task specific (78). The generalized effects on gross motor development may reflect the whole-body nature of crawling, which not only stimulates the central and peripheral nervous system but also engages a large percentage of the infant’s musculature.

### Effect of the Crawling Intervention on General Development

The broader effects of crawling training on the total ASQ-3 score, and specifically on the scores representing the communication domain, is likely a result of the additional experiences associated with moving through space and their perceptual consequences. These experiences occurred during the eight weeks of training and as a function of the faster acquisition of crawling in the Crawli group. As noted in the introduction, considerable research has linked locomotor development to developmental changes in perception-action coupling, spatial cognition, memory, and social and emotional functioning during the first year of life (6,17,79) and to language development at 12 months of age (8,28). Researchers have speculated that independent locomotion allows infants to have new experiences which in turn recruit the processes that drive developmental change in domains far removed from the locomotor domain (6,17). Taken together, these findings highlight that an intervention focused on training a specific motor skill, crawling, can have broad effects on the motor domain of functioning as well as on other domains of functioning seemingly unconnected to the motor domain.

### Possible mechanisms underlying the effects of the early crawling intervention

The fact that Crawli but not Mattress training facilitates the development of mature crawling permits different hypotheses. Although when infants lie prone on a mattress they cannot propel themselves very far, they make numerous leg movements (not arm movements), which are inefficient but present. This observation suggests that one of the central components of the “Crawli effect” is propulsion and/or the ability to perform coordinated quadrupedal leg and arm movements. It is impossible to list here the many possible mechanisms behind the effects of Crawli training but some would be especially interesting to investigate in the future. These include the effect of training on the morphology and composition of the leg and arm muscles and on the quadrupedal inter-limb coordination and the intra-limb coordination. It would also be interesting to investigate the evolution of muscle synergies using electromyography in relation to training. Another mechanism likely playing an important role in the effect of Crawli training is the stimulation of perception-action loops between the propulsion actively generated by the infant and the resulting proprioceptive, visual and auditory consequences associated with movement through the environment. We intentionally restricted motivational and visual/auditory stimuli in this study during training to minimize potential bias. However, it would be particularly interesting to add visual, auditory, and/or olfactory stimuli during clinical trainings given prior research showing that these stimuli can facilitate newborn stepping and crawling (50–54), especially if these stimuli were combined with parental encouragements.

### Limitations

The main limitation of this intervention is the restricted number of participants, which was a function of the number of osteopaths needed to supervise the large number of home training sessions. However, this design feature was necessary to ensure the training protocol was administered faithfully and to monitor for any possible adverse effects of the training. The small sample size meant that it was not possible to do subgroup analyses within the Crawli group to see, for example, whether any individual difference characteristics explained why some infants derived more benefit from the intervention than others or to see if infants who traveled greater distances during training also had better developmental outcomes. Another limitation of the intervention was the decision to exclude infants with major brain lesions. However, the success of the current intervention suggests the Crawli intervention should now be tested on infants with brain damage. A final design limitation was the daily training time limited to 5 minutes and the decision to end training after 8 weeks. A longer duration of training may have further increased the efficacy of the intervention.

## CONCLUSION

Our early training in crawling intervention facilitated the acquisition of mature independent crawling and had broad effects on development, which were evident from 2 to 12 months of age, but were strongest at two and six months of age. With a longer training duration, the effects might have been even stronger. The feasibility and efficacy of our intervention for very premature infants without major cerebral lesions opens new opportunities to apply this method to a larger number of infants and to infants with brain lesions. Training early crawling on a Crawliskate is a relatively inexpensive, simple, and effective way to stimulate premature infants’ motor and general development during the first year of life.

## Data Availability

The data that support the findings of this study are available on request from the corresponding author. The data are not publicly available due to privacy or ethical restrictions.

## Acknowledgements

This work was supported by the grant ANR-20-CE17-0014 from the Agence Nationale de la Recherche, the grant Project-106 from the SATT-Erganeo (Société d’Accélération des Transferts de Technologies) and a fellowship (ED261) from the French Government to support Marie-Victorine Dumuids-Vernet’s doctoral thesis. We would like to thank all the infants and parents who participated in this study and all the osteopaths who performed the 1600 total home visits for their enthusiastic support during this study.

## VIDEO LEGENDS

Note: videos of human subjects are not posted on MedRxiv, but are available under request to the corresponding author.

**Video Crawli Home.** Video of a premature girl experiencing her first session on the Crawliskate at home. While this is her first experience with the Crawliskate, the infant is able to propel herself in a short time. The beginning of the video shows the osteopath helping the infant by repositioning her feet and arms on the mattress.

**Video Mat Home.** Video of a premature infant prone on the Mattress without the help from the Crawliskate. While the infant preforms many leg movements, arm movements are mostly blocked and propulsion is impeded.

**Video Crawli Lab.** The video shows a test on the Crawliskate at our babylab of a premature infant just discharged from the NICU before the home training started. His tolerance for the Crawliskate was evaluated by recording his oxygen saturation and heart rate.

**Video Mat Lab.** The video shows a test on the Mattress at our babylab of a premature infant just discharged from the NICU before the home training started.

## Footnotes

1 Photos (Figure 2) and videos of human subjects are not posted on MedRxiv, but are available under request to the corresponding author.

2 Note, we do not include the Bayley III data at 24 months or the ASQ-3 at 18, 24 and 60 months in this report as we plan to publish those data in a later report on the long-term effects of the intervention.

